# Determining county-level counterfactuals for evaluation of population health interventions: A novel application of *K*-means cluster analysis

**DOI:** 10.1101/2020.04.30.20086124

**Authors:** Kelly L. Strutz, Zhehui Luo, Jennifer E. Raffo, Cristian I. Meghea, Peggy Vander Meulen, Lee Anne Roman

## Abstract

**Objectives:** Evaluating population health initiatives at the community level necessitates valid counterfactual communities, which includes having similar complexity with respect to population composition, healthcare access, and health determinants. Estimating appropriate county counterfactuals is challenging in states with large inter-county variation. We present and discuss an application of *K*-means cluster analysis for determining county-level counterfactuals in an evaluation of a county perinatal system of care for Medicaid-insured pregnant women.

**Materials and Methods:** Counties were described using indicators from the American Community Survey, Area Health Resources Files, University of Wisconsin Population Health Institute County Health Rankings, and vital records for Michigan Medicaid-insured births for the year intervention began (or the closest available year). We ran analyses of 1,000 iterations with random starting cluster values for each of a range of number of clusters from 3 to 10 and used standard variability and reliability measures to identify the optimal number of clusters.

**Results:** One county was grouped with the intervention county in all solutions for all iterations and thus considered most valid for 1:1 population county comparisons. Two additional counties were frequently grouped with the intervention county. However, no county was ideal for all subpopulation analyses.

**Practice Implications:** Although the K-means method was successful at identifying a comparison county, concerning intervention-comparison differences remained. This limitation of the method may be specific to this county and the constraints of a within-state study. This method could potentially be more useful when applied to other counties in and outside of Michigan.

## Introduction

Complex, multifactorial public health problems necessitate population-wide solutions, and thus community-level interventions are frequently utilized. The gold standard for evaluating community-level interventions, such as those at the county level, is the group-randomized trial.^1^ However, this is often not feasible or appropriate with a small number of communities, particularly for community-based participatory research and other scenarios where the intervention is community- or stakeholder-initiated, or system of care interventions that depend on coordinating local resources. It is not clear how best to evaluate non-randomly assigned community interventions in this scenario.^2–4^ As in individual-level quasi-experimental analyses, the challenge is to identify an appropriate counterfactual group or communities that should be as similar as possible to the intervention community/ies to approximate group randomization,^3,5^ but little guidance exists on selection criteria or methods.

We conducted a demonstration project from 2009-2015 to determine whether a county-level perinatal system of care enhancement improved population outcomes for Medicaid-insured pregnant women and their infants in Kent County, Michigan, a mixed urban/rural county containing the second-largest city in the state. Because our intervention targeted the entire county, the ideal counterfactual community or communities would have the same sociodemographic characteristics and level of complexity in service delivery as the intervention county. Complexity is influenced by many factors, including the numbers of residents, health care agencies, providers, and enhanced prenatal care programs.

Because we wanted to account for the broader context and administrative features of an entire county, we explored a clustering method, *K*-means cluster analysis, to identify one or more comparison counties in Michigan. *K*-means cluster analysis has been used increasingly in the public health and health services research literature to create clusters of exposures or individuals, most frequently for sources of air pollution^6^ and for health care population segmentation.^7^ Where it has been used previously for county-level clustering, it was employed to create clusters of similar counties to compare among each other.^8,9^

The objective of this study was to present and discuss an application of *K*-means cluster analysis for determining county-level counterfactuals for evaluation of a county perinatal system of care for Medicaid-insured pregnant women. To our knowledge, this is the first use of this method to find the most similar counterfactual county.

## Materials and Methods

### Data

There are 83 counties in the state of Michigan. County-level features were selected from three publicly available data sources – the American Community Survey,^10^ the Area Health Resources Files,^11^ and the University of Wisconsin Population Health Institute County Health Rankings.^12^ The fourth data source was a limited dataset of Michigan resident live birth records for Medicaid-insured births only (hereafter called Michigan Medicaid vital records (MMVR)), including county identifiers. The file came from the Michigan Department of Health and Human Services (MDHHS) Division for Vital Records and Health Statistics, and was retrieved by an honest broker from the MDHHS Health Services Data Warehouse.

The following categories of features were considered: population composition, social and economic factors, health care access, health outcomes, health behaviors, and physical environment. We selected a total of 35 features across these broad categories, some chosen from Rettenmaier and Wang^13^ and others based on the evaluation team’s experience with the Michigan Medicaid Maternal and Infant Health Program and the Strong Beginnings federal Healthy Start program.^14–16^ These represented a mixture of compositional (i.e., those aggregated from individual-level characteristics) and contextual (i.e., those pertaining to the county environment) measures.^17^ Whenever possible, we used year 2009 data because it was the year when system implementation of pregnancy-centered service improvement began. For data elements without 2009 information, we used as proxies, in priority order, 2010, 2008, or 2011 data. Four features were missing data for certain counties (n ranged from 14-35) and mean values were imputed. A description of the 35 features utilized is in Table 1 along with the data source, year, mean, and standard deviation.

**Table 1.**
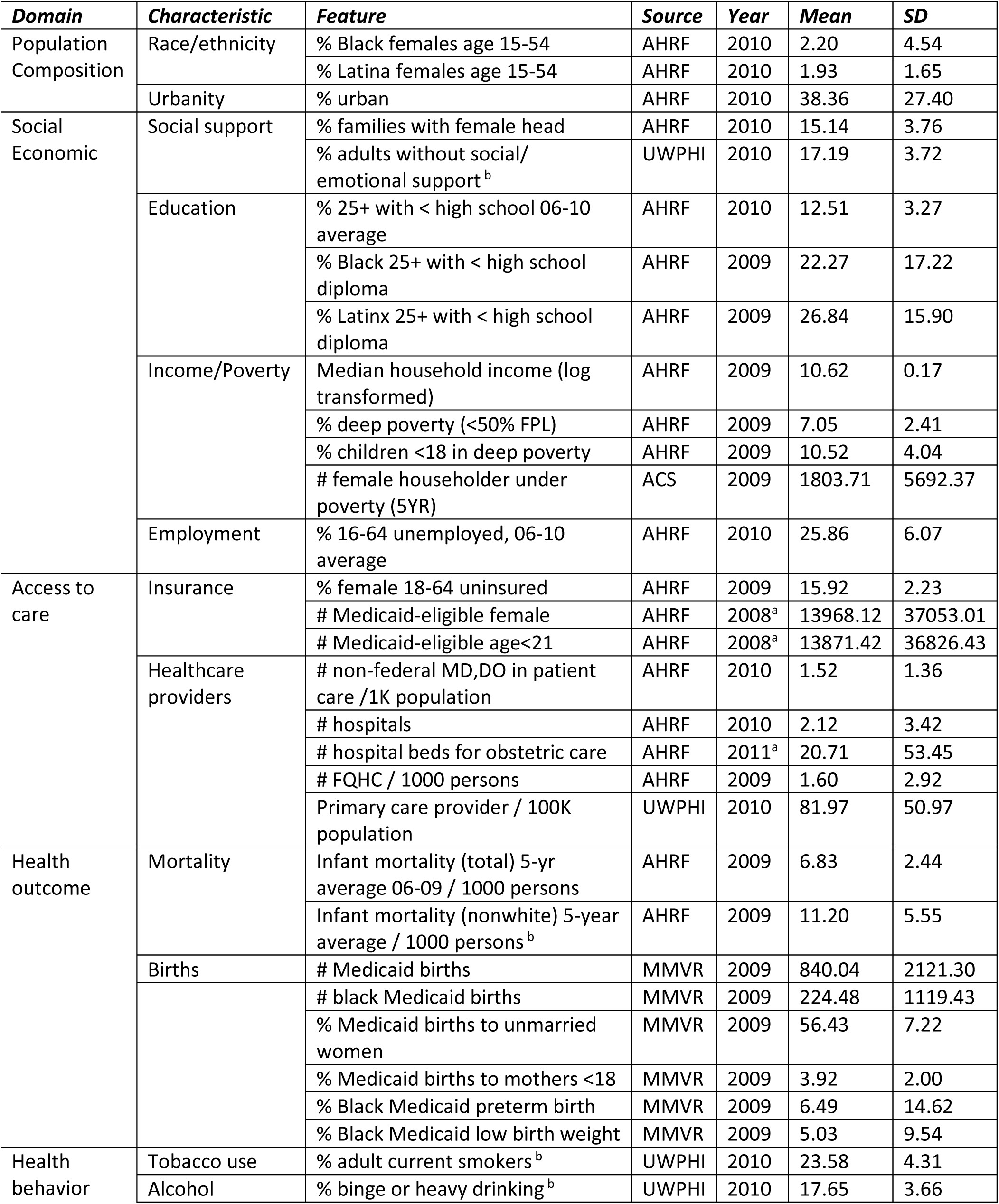
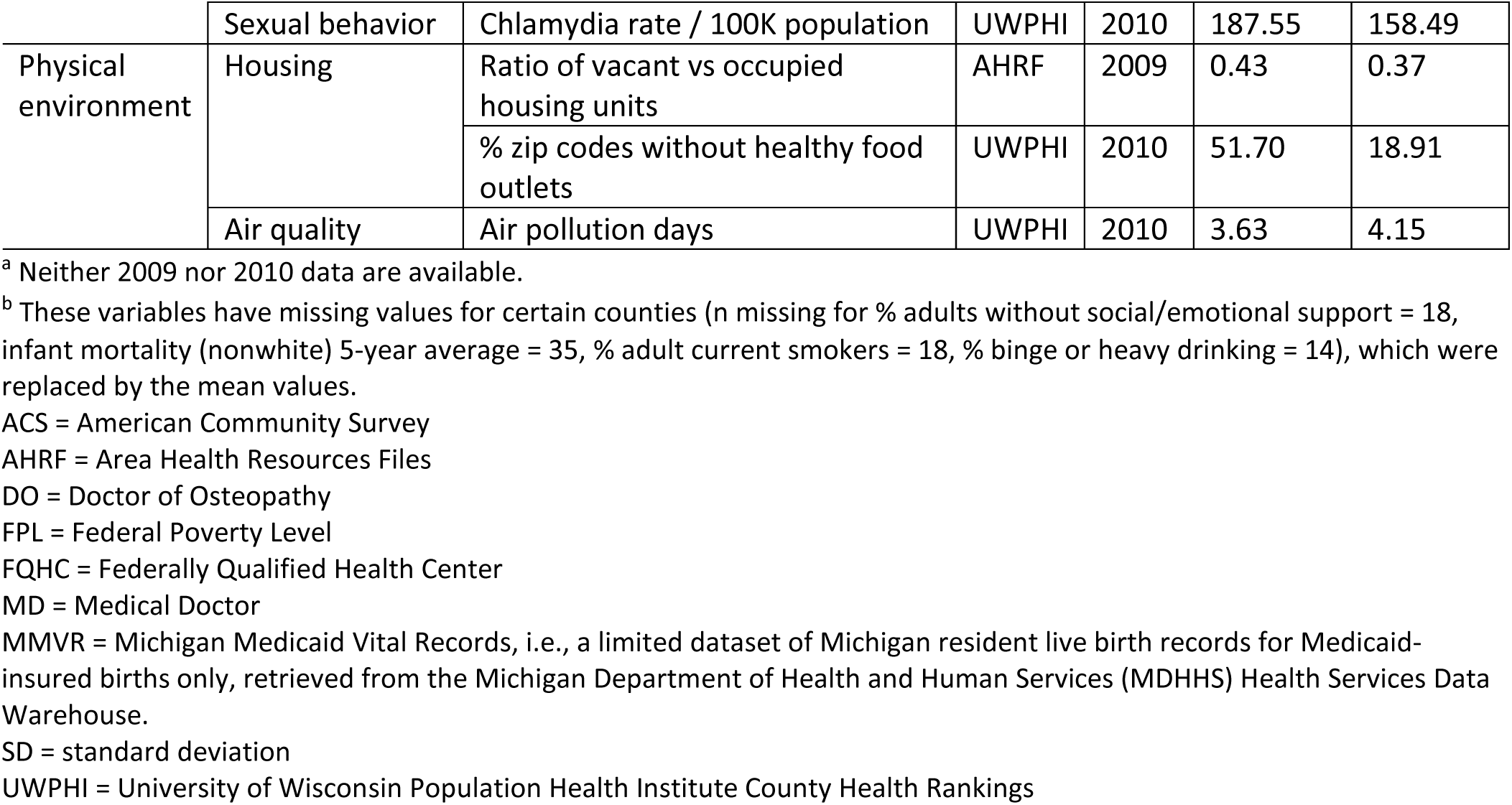
County-level features used to characterize Michigan counties from the American Community Survey, Area Health Resources Files, Michigan Medicaid Vital Records, and University of Wisconsin Population Health Institute County Health Rankings, 2008-2011.

### Analysis

*K*-means cluster analyses were used to identify the comparison county or counties that would be classified in the same cluster as the intervention county in the most optimal model. Cluster analysis, also called data segmentation, groups or segments a collection of objects (counties in our study) into subgroups or clusters such that those within the same cluster are more similar to each other than objects (counties) assigned to different clusters.^18^ Counties are described by a set of characteristics (features, measurements) that are pre-selected based on the purpose of evaluations; in our case, relevant to evaluations of pregnancy-centered health programs. Counties in the same cluster may have different degrees of similarity or dissimilarity to each other and to a specific county (Kent in our study). Among the most widely used clustering methods is the K-means (or median) clustering for which the within-cluster variation (WCV) is as small as possible.^6,7^

We tested solutions for 3 to 10 clusters for 1,000 iterations each with randomly assigned starting cluster values. To identify the best solution, we used scree plots to visualize a kink in the curve generated from the within sum of squares (WSS) or its logarithm (log[WSS]) for all cluster solutions, i.e., the point at which reduction in WSS or logWSS is not appreciable enough to warrant increasing the number of clusters.^19^ The WSS can be thought of as the error sum of squares in regression analysis where the total sum of squares is defined when all objects are in one cluster. Another criterion used was the η^2^ coefficient, 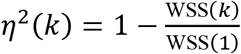, which is between 0 and 1 with higher values indicating better clustering. Finally, we also transformed the WSS to get the proportional reduction of error (PRE) as well to identify the number of clusters that leads to the largest reduction in PRE.

Nineteen of the 35 features are proportions which have a natural range (0 and 100); and the other features may have large variation. It was conceivable that if we standardized these features some natural variation, e.g., the size of the population in each county, would be masked. In *K*-means analyses, if all variables are standardized then clustering based on correlation (similarity) is equivalent to that based on squared distance (dissimilarity).^20^ Therefore, we ran all analyses twice, with and without standardizing all variables. Stata v.15 (StataCorp, College Station, TX) was used for all analyses. The evaluation project was reviewed by the Michigan State University, MDHHS, and Spectrum Health institutional review boards and determined not human subject research.

## Results

Figure 1 shows that the optimal number of clusters when features were not standardized was 4, where WSS could be seen at a kink point and the reduction in PRE was the largest. On the other hand, Figure 2 shows that when features were standardized there was no clear optimal solution of the number of clusters.

**Figure 1.**
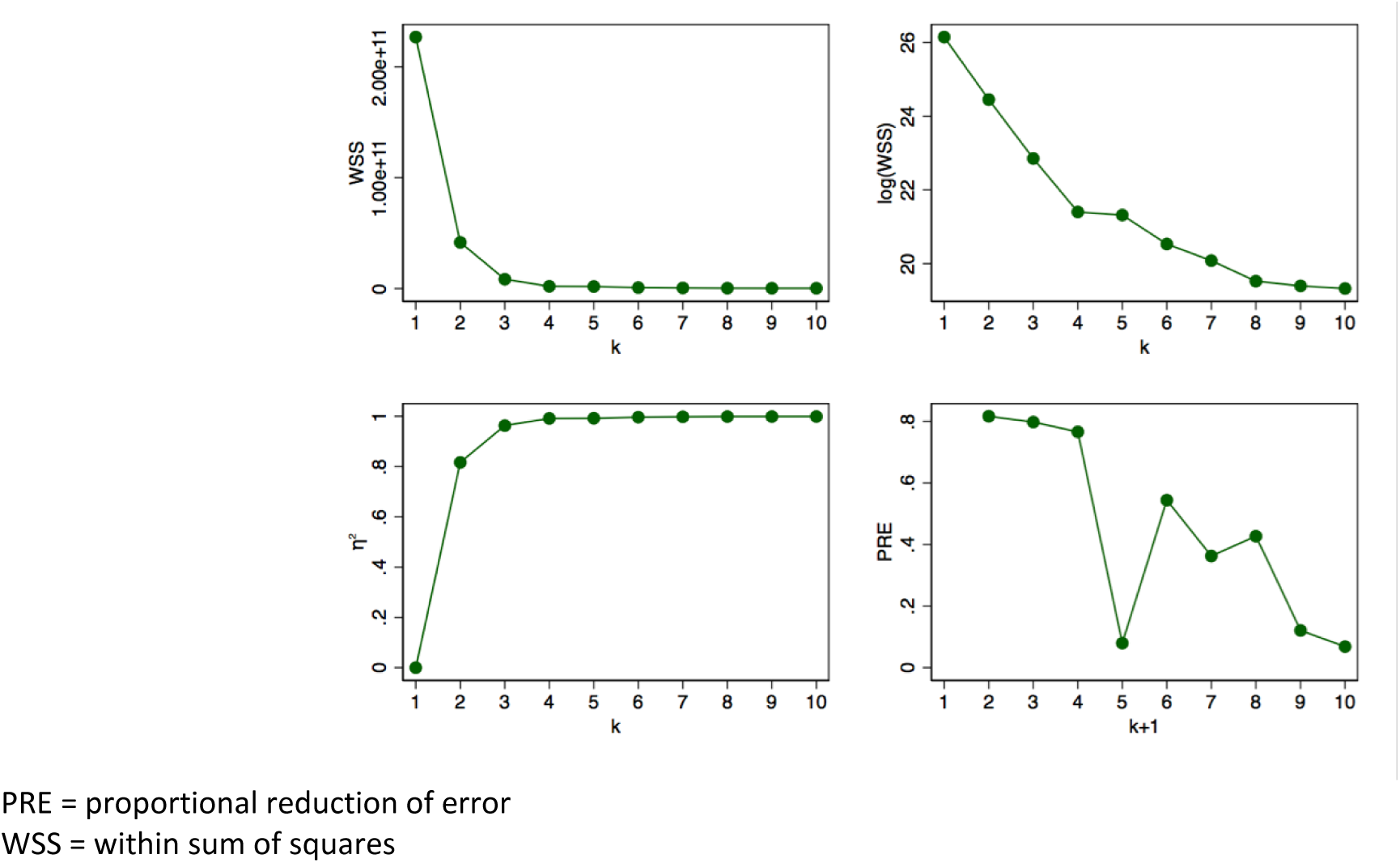
WSS, log(WSS), *η*^2^ coefficient and PRE for 10 cluster solutions when features are not standardized, Michigan, 2008-2011.

**Figure 2.**
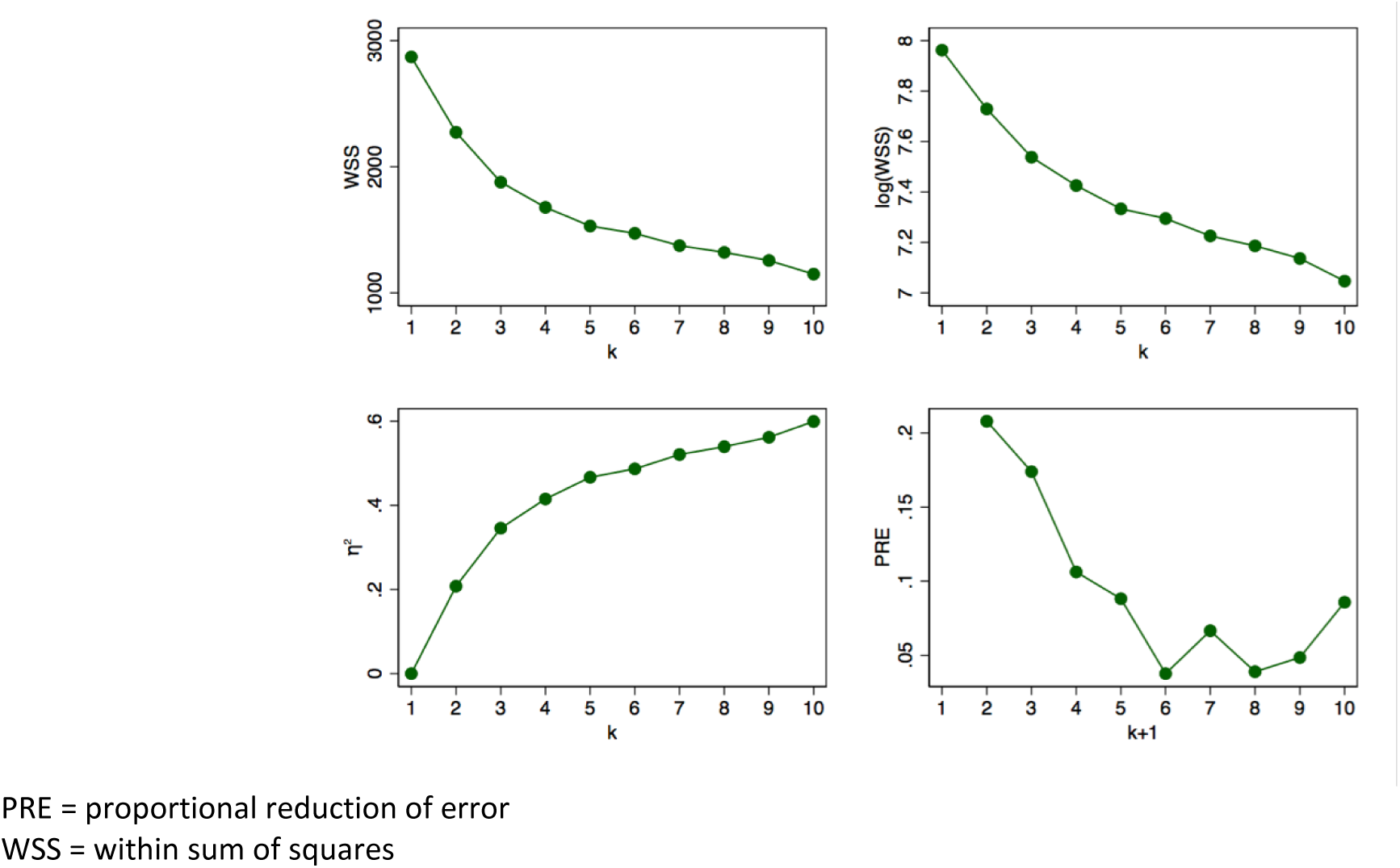
WSS, log(WSS), *η*^2^ coefficient and PRE for 10 cluster solutions when features are standardized, Michigan, 2008-2011.

Using the unstandardized features, the number of times that a county was grouped in the same cluster as Kent County out of 1,000 iterations when the number of clusters varied from 3 to 6, the range around the optimal number of 4, is shown in Table 2. Three counties were grouped with Kent under these criteria. Macomb County was grouped in the same cluster with Kent County in all iterations of all solutions, while Oakland County clustered with Kent in 979-1000 iterations and Genesee County clustered in 127-1000 iterations depending on number of clusters. In contrast, when we standardized all features, no county was grouped with Kent in a majority of iterations (results available upon request). Given the lack of an optimal solution under standardization, and that the counties grouped together were not similar, we opted to use the unstandardized variables.

**Table 2.**
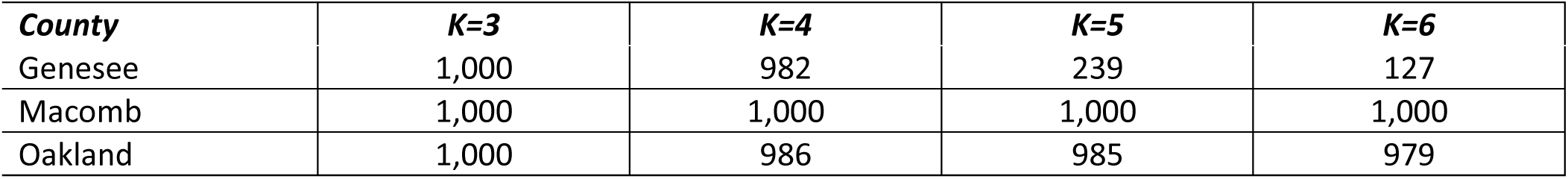
Number of times a county is grouped together with Kent County in 1,000 iterations, Michigan, 2008-2011.

| ***County*** | ***K=3*** | ***K=4*** | ***K=5*** | ***K=6*** |
| --- | --- | --- | --- | --- |
| Genesee | 1,000 | 982 | 239 | 127 |
| Macomb | 1,000 | 1,000 | 1,000 | 1,000 |
| Oakland | 1,000 | 986 | 985 | 979 |

Table 3 lists the values of the 35 features for Kent, Macomb, Oakland, and Genesee counties. In examining the values individually, Macomb was within 2 percentage points of Kent for 10 of 19 percentage-based features, with nearly identical median household income and numbers of Medicaid-eligible women and minors. Oakland and Genesee were within 2 percentage points of Kent for 5 of 19 percentage-based features each, also both with similar median household incomes. There was broad variation in access to care features across counties. However, there were two related features for which all of the three comparison counties had values considerably smaller than Kent’s: the two Latinx population features, “percent Latina females ages 15-54” (Kent 6.7, Macomb 1.5, Oakland 2.4, Genesee 2.0) and “percent Latinx aged 25+ with less than a high school diploma” (Kent 46.3, Macomb 22.4, Oakland 27.0, Genesee 16.9).

**Table 3.** Features for Kent, Macomb, Oakland, and Genesee Counties, Michigan, 2008-2011.

| **Feature** | **Kent** | **Macomb** | **Oakland** | **Genesee** |
| --- | --- | --- | --- | --- |
| % Black females age 15-54 | 7.3 | 7.0 | 10.5 | 14.9 |
| % Latina females age 15-54 | 6.7 | 1.5 | 2.4 | 2.0 |
| % urban | 84.3 | 97.2 | 95.2 | 83.2 |
| % families with female head | 18.8 | 18.9 | 16.9 | 26.0 |
| % adults without social/emotional support | 18.2 | 20.2 | 17.3 | 23.0 |
| % 25+ with < high school 06-10 average | 11.7 | 12.4 | 7.8 | 11.9 |
| % Black 25+ with < high school diploma | 19.7 | 13.5 | 10.3 | 16.3 |
| % Latinx 25+ with < high school diploma | 46.3 | 22.4 | 27.0 | 16.9 |
| Median household income (log transformed) | 10.8 | 10.8 | 11.0 | 10.6 |
| % deep poverty (<50% FPL) | 6.8 | 4.8 | 4.3 | 9.5 |
| % children <18 in deep poverty | 9.6 | 7.0 | 5.7 | 15.5 |
| # female householder under poverty (5YR) | 8864.0 | 7333.0 | 9024.0 | 9745.0 |
| % 16-64 unemployed, 06-10 average | 19.5 | 22.7 | 20.8 | 30.7 |
| % female 18-64 uninsured | 14.8 | 14.7 | 12.3 | 13.0 |
| # Medicaid-eligible female | 72009.0 | 74763.0 | 79324.0 | 65315.0 |
| # Medicaid-eligible age<21 | 74313.0 | 73401.0 | 78119.0 | 65053.0 |
| # non-federal MD,DO in patient care /1K population | 3.3 | 1.6 | 5.7 | 2.5 |
| # hospitals | 7.0 | 8.0 | 17.0 | 4.0 |
| # hospital beds for obstetric care | 177.0 | 55.0 | 255.0 | 90.0 |
| # FQHC / 1000 persons | 11.0 | 1.0 | 2.0 | 5.0 |
| Primary care provider / 100K population | 129.3 | 71.0 | 207.5 | 132.1 |
| Infant mortality (total) 5-yr average 06-09 /1K population | 7.4 | 6.5 | 6.4 | 9.2 |
| Infant mortality (nonwhite) 5-year average /1K population | 13.7 | 10.7 | 10.8 | 16.9 |
| # Medicaid births | 5045.0 | 4830.0 | 5320.0 | 3739.0 |
| # black Medicaid births | 1005.0 | 947.0 | 1506.0 | 1407.0 |
| % Medicaid births to unmarried women | 60.4 | 52.8 | 57.3 | 71.8 |
| % Medicaid births to mothers <18 | 4.7 | 2.9 | 3.5 | 6.3 |
| % Black Medicaid preterm birth | 13.5 | 12.8 | 14.3 | 17.0 |
| % Black Medicaid low birth weight | 12.1 | 13.2 | 12.7 | 15.2 |
| % adult current smokers | 21.3 | 22.5 | 18.4 | 25.9 |
| % binge or heavy drinking | 17.1 | 18.6 | 16.0 | 16.0 |
| Chlamydia rate / 100K population | 559.9 | 195.8 | 274.7 | 709.3 |
| Ratio of vacant vs occupied housing units | 0.1 | 0.1 | 0.1 | 0.1 |
| % zip codes without healthy food outlets | 55.1 | 14.6 | 33.3 | 45.9 |
| Air pollution days | 15.0 | 9.0 | 10.0 | 8.0 |

DO = Doctor of Osteopathy
FPL = Federal Poverty Level
FQHC = Federally Qualified Health Center
MD = Medical Doctor

## Discussion

*K*-means clustering achieved identification of one to three comparison counties when pre-selected features were unstandardized in our study. Yet, the features that differed between the intervention county and other similar counties were concerning. In particular, the intervention county had more Latina women, and more with less than a high school education, than the best matched county by the *K*-means analysis. We were also cautious about differences in county maternal and infant health programming and infrastructure that were not reflected in the data sources. For example, one of the selected counties includes the city of Flint that would receive an influx of maternal and child health resources associated with the water crisis during the study period. Ultimately, we were able to achieve better covariate balance by selecting individual women from across the state (outside the intervention county) and using propensity score-weighted analyses, matched exactly on race and using more granular census tract-level and block group-level covariates.

These results are consistent with the findings of Wallace et al.,^8^ who used *K*-means clustering with national U.S. data to create clusters of counties similar on sociodemographic features. They noted considerable within-state heterogeneity, such that most states (including Michigan) contained counties from many different clusters. Of note, the three comparison counties that we identified were in the same cluster as our intervention county in Wallace’s schema as well. However, they also identified that the counties that were themselves sociodemographically heterogenous would benefit from more granular, “subcounty” data, which we found applicable to our diverse intervention county.

Our findings are also congruent with recent discussion of how machine learning algorithms can reproduce biases against minoritized groups.^21–23^ While previous literature has focused on racial/ethnic disparities in predictive algorithms, our analysis demonstrates how an ethnic subgroup can be obscured by a clustering method. Thus, the same cautions that should be extended to ensure equity in predictive modeling should be applied to cluster analyses as well.

General limitations of the *K*-means method to select counterfactual communities include the following. First, the results are only as good as the chosen features. In our example, features were selected through discussion by the research team and community partners with substantive knowledge of factors influencing perinatal health in the community. We were able to capture a wide range of important indicators of complexity, including health care determinants and social determinants of maternal and infant health outcomes, triangulated across four different data sources. That said, our selection was likely not the optimal set, and it is probable that no one optimal set exists. A more data-driven approach could be applied to feature selection as well.^8^ Relatedly, given our interest in the Latinx subpopulation, adding more measures specific to this subpopulation would weight their importance more heavily. However, this would not address the issue that there aren’t appreciable Latinx subpopulations in the counties whose other features are most similar to our intervention county, and would have led to no better solution.

Second, this method involves the assumption that if a counterfactual community is comparable at baseline, it will remain so throughout the study. This may not be the case; changes may occur during the intervention period that make a community more or less comparable to the intervention community.^3^ This limitation is shared with any method for identifying a comparison at baseline. As noted above, this became particularly salient to our example given the Flint water crisis and resulting resource allocation. Future research is needed to adapt these methods for changes across time. Moreover, in our data sources, not all features were measured during the optimal baseline year, necessitating the use of proxy years. However, the intervention to be evaluated was complex and involved many changes over the years, and even the post-baseline proxy years were from before many of the changes were implemented.

Finally, the use of *K*-means clustering in general comes with its own set of assumptions and limitations.^20^ One issue of the *K*-means clustering method is that the resulting assignments depend on the random starting point. The *K*-means algorithm gives local minima and does not guarantee to give the global minimum, so the starting points should be varied to examine the end partitioning. Another issue of the *K*-means algorithm is that a variable with high variability can dominate the cluster analysis. A common solution is to standardize variables, but standardizing could also hide the true groupings in the data. This is a case-by-case decision depending on the type of data and the nature of the groups. The number of clusters *K* is the tuning parameter that can be chosen by cross-validation if there are large number of independent and identically distributed observations, which was not possible in our presented example.

## Practice Implications

*K*-means clustering provides a rigorous, data-driven approach to selecting one or more counterfactual groups for a community-level intervention, representing an improvement over subjective selection of comparison communities. Although it provided a consistent comparison county solution, it did not result in an optimal comparison for the intervention county. The method would be potentially more useful at other geographic levels or for other counties in or outside of Michigan. Nonrandomly assigned community interventions are common in population health; the present analysis can stimulate conversation about how best to select appropriate comparisons. In particular, it demonstrates that selection of counterfactual communities should be objective, transparent, and examined critically using a health equity lens.

## Data Availability

Three of four data sources (the American Community Survey, the Area Health Resources Files, and the University of Wisconsin Population Health Institute County Health Rankings) are publicly available at the links below. The fourth was obtained under a data use agreement from the Michigan Department of Health and Human Services (MDHHS) Health Services Data Warehouse; interested parties would need to contact MDHHS.

https://www.census.gov/programs-surveys/acs/

https://data.hrsa.gov/topics/health-workforce/ahrf

http://www.countyhealthrankings.org

## Acknowledgements

The authors thank the Michigan Department of Health and Human Services for access to their Health Services Data Warehouse and consultation from the Maternal and Infant Health Division and the Division for Vital Records and Health Statistics.

